# Maternal obesogenic environment and its association with childhood obesity in Peru: A 9-year analysis of a national survey

**DOI:** 10.1101/2024.08.11.24311822

**Authors:** Joan A. Loayza-Castro, Víctor Juan Vera-Ponce, Luisa Erika Milagros Vásquez-Romero, Jhonatan Roberto Astucuri Hidalgo, Nataly Mayely Sanchez-Tamay, Fiorella E. Zuzunaga-Montoya

## Abstract

**Introduction:** Childhood obesity is a global public health concern with increasing prevalence in Peru. The obesogenic environment, including maternal and family environmental factors, plays a crucial role in the development of childhood obesity.

**Objective:** To analyze the association between the maternal obesogenic environment and obesity in children under five years of age in Peru.

**Methods:** An analytical cross-sectional study used data from the Demographic and Family Health Survey (DHS) from 2014 to 2022. To assess the obesogenic environment, variables such as maternal obesity, television use, smoking, and maternal anemia were analyzed. Childhood obesity was a body mass index Z-score > +3 standard deviations. A Poisson regression model was used to calculate crude and adjusted prevalence ratios.

**Results:** The prevalence of childhood obesity was 1.99%. Obese mothers were found to be 1.52 times more likely to have obese children (aPR=1.52, 95% CI 1.40-1.65; p<0.001). No significant associations were found between frequent television use, maternal smoking, and anemia with childhood obesity after adjusting for multiple factors.

**Conclusion:** This study highlights the importance of the maternal obesogenic environment, especially maternal obesity, in developing childhood obesity in Peru. Comprehensive interventions that address multiple aspects of the family obesogenic environment, including the prevention and management of maternal obesity, promotion of healthy lifestyles, and strengthening of public policies that foster healthy environments, are recommended.

## Introduction

Childhood obesity currently represents a global public health problem, characterized by a Body Mass Index (BMI) above average for the child’s age and sex. According to the World Health Organization (WHO), the prevalence of childhood obesity has increased alarmingly, affecting more than 40 million children under five years of age worldwide in 2022 ^(1)^. In Peru, according to the latest National Demographic and Family Health Survey (DHS) of 2023, 19.4% of children under five years old are overweight or obese, representing a significant increase compared to previous years ^(2)^.

The concept of the obesogenic environment is crucial to understanding the obesity epidemic. This term refers to environmental conditions that promote weight gain and obesity, including factors such as the availability and accessibility of unhealthy foods, lack of spaces for physical activity, and social and cultural influences that encourage unhealthy eating habits ^(3)^. In the family context, the maternal obesogenic environment plays a fundamental role in the development of childhood obesity ^(4)^.

Maternal obesity, as an integral part of the family obesogenic environment, is intrinsically related to the risk of childhood obesity. Recent studies have shown that maternal obesity significantly increases the likelihood of their children developing obesity, both at birth and throughout their lives ^(5)^. Globally, the prevalence of maternal obesity is alarming, with more than 25% of women of reproductive age being overweight or obese ^(6)^.

However, the maternal obesogenic environment goes beyond obesity per se. Factors such as sedentary habits, represented by excessive television use ^(7)^, maternal smoking ^(8)^, and the nutritional quality of the home, indirectly reflected by the presence of anemia in the mother ^(9)^, contribute to creating an environment conducive to the development of childhood obesity, thus shaping the obesogenic environment.

The influence of the maternal obesogenic environment on the development of childhood obesity is an emerging and crucial area of research. However, in Peru, despite the growing concern about childhood obesity, few studies have examined in depth the relationship between the maternal obesogenic environment. This knowledge gap is significant given the rapid change in lifestyles and dietary patterns that the country has experienced in recent decades.

Therefore, this study aims to analyze the association between the maternal obesogenic environment, including maternal obesity, and obesity in children under five in Peru, using data from the DHS from 2014 to 2022. Specifically, we examine how factors such as maternal obesity, television use, smoking, and the presence of anemia relate to the prevalence of childhood obesity.

The results of this study could provide valuable information for developing interventions and public policies aimed at preventing childhood obesity in Peru. These policies should address not only individual factors but also environmental and family factors contributing to this growing epidemic.

## Materials and Methods

### Study Design and Context

This study employed an analytical cross-sectional design based on data from the Peruvian Demographic and Family Health Survey, conducted between 2014 and 2022. DHS is a nationally representative annual survey that collects information on the Peruvian population’s health, nutrition, and demographics.

### Population, Selection Criteria, and Sample

The study population included mothers and children under five who participated in DHS between 2014 and 2022. DHS utilizes a probabilistic, stratified, two-stage sampling method, differentiating between urban and rural areas, allowing for a representative sample at national and departmental levels ^(10)^.

The inclusion criteria were mothers aged 12 to 49 with at least one child under 5. The exclusion criteria included incomplete weight and height data for both mother and child.

### Variables and Measurement

The primary dependent variable was childhood obesity, assessed using the body mass index for age (BMI/age) according to World Health Organization (WHO) growth standards for children under five years. A child was considered obese when the BMI Z-score exceeded +3 standard deviations ^(11)^.

Key independent variables included maternal obesity, evaluated through BMI, with obesity classified as BMI ≥ 30 kg/m² ^(1)^. Television use was categorized into three levels based on reported frequency in DHS: never, occasional (less than once a week), and frequent (at least once a week). Maternal smoking status was dichotomized as non-smoker or smoker, with smokers defined as those reporting current tobacco use, regardless of frequency.

Maternal anemia was determined from hemoglobin levels adjusted for altitude, using WHO cut-off points, with anemia defined as hemoglobin levels < 12 g/dL ^(12)^.

Various maternal sociodemographic variables were included to control for potential confounding factors. Maternal age (in years), maternal sex, educational level (categorized as no education, primary, secondary, and higher), marital status (dichotomized as single or partnered), natural region of residence (categorized as Metropolitan Lima, rest of the coast, highlands, and jungle), and area of residence (urban or rural) were considered. For children, in addition to obesity, two additional sociodemographic variables were considered: the child’s age (categorized as less than two years and 3 to 5 years) and the child’s gender (female or male).

All variables were obtained directly from the DHS database, using definitions and categorizations provided by Peru’s National Institute of Statistics and Informatics, except for childhood obesity and maternal anemia, which were calculated using the mentioned international standards.

## Procedures

Data from DHS for 2014 to 2022 were accessed through the INEI web portal, which provides public access to this information.

Both child and maternal obesity were calculated using weight and height data in the DHS anthropometry module. BMI was calculated by dividing the individual’s weight (in kilograms) by their height (in square meters). Weight was measured to the nearest 0.1 kg using an electronic scale, with the individual wearing light clothing and no shoes. Height was measured to the nearest 0.1 cm using a wooden stadiometer, following international standards and Peruvian guidelines.

Television use was assessed using information from the household characteristics module, analyzing the question: “What is the frequency of watching television?” Maternal smoking status was obtained from the women’s health module, using the question on current tobacco consumption: “Do you currently smoke?”

For hemoglobin testing, participants were informed that the test would involve extracting a small drop of blood from their fingers using guaranteed and safe equipment and disposable supplies designed specifically for each person. They were assured that results would be communicated immediately and handled confidentially.

Sociodemographic variables such as maternal age, educational level, marital status, and area of residence were extracted from the household and interviewee characteristics module. The natural region was determined from the geographical information provided in the DHS database.

Age and sex were obtained from the birth history module for children. Age was calculated using the date of birth and interview date, while sex was taken directly from the provided information.

### Statistical Analysis

This research used the statistical package R version 4.3.1. The analysis was conducted in several stages.

First, a descriptive analysis of all study variables was performed. For categorical variables, absolute and relative frequencies were calculated and presented as percentages. Functions from the ‘summarytools’ package in R were used to generate these descriptive statistics.

Additionally, a graphical analysis was performed to visualize relationships between the main factors of the maternal obesogenic environment and child BMI. Scatter plots were generated to examine correlations between maternal BMI and child BMI, as well as between maternal hemoglobin levels and child BMI. Box plots were created to visualize the distribution of child BMI according to television use and maternal smoking status. These graphical analyses were performed using the ggplot2 package.

Subsequently, a Poisson regression model with robust variance was used to calculate crude (cPR) and adjusted (aPR) prevalence ratios with their 95% confidence intervals (95% CI).

It is important to note that all analyses were performed considering the complex design of the DHS survey, using the provided sampling weights. R’s ‘survey’ package incorporated the survey design into the analyses.

### Ethical Aspects

This study strictly adhered to the ethical principles established in the Declaration of Helsinki and good research practice standards. Although secondary data from publicly accessible DHS were used, measures were taken to ensure the information was handled ethically.

Due to the secondary and anonymous nature of the data used, this study did not require specific approval from an ethics committee. However, it was verified that DHS, the data source, had obtained the corresponding ethical authorization for data collection.

## Results

Regarding the nutritional status of children, the prevalence of obesity was 1.99%, while for mothers it was 23.07%. For the maternal obesogenic environment, television use was considered, where 59.18% reported frequent use and smoking activity, and 2.35% of participants indicated they were smokers.

Considering the sociodemographic variables of the mothers who participated in the study, 40.57% were between 20 and 29 years old, 65.95% reported having secondary education, and 72.08% had a partner. On the other hand, 27.61% of the participants lived in the jungle, 73.84% lived in urban areas, and 21.48% had anemia. Regarding the children, the findings were that 50.91% were male, and 75.14% were less than two years old.

**Table 1.** Characteristics of participating mothers and children.

| **Characteristic** | **n = 55,190** |
| --- | --- |
| **Age group** |  |
| 12 - 20 years | 7,632 (13.83%) |
| 20 - 29 years | 22,391 (40.57%) |
| 30 - 39 years | 19,590 (35.50%) |
| 40 - 49 years | 5,577 (10.10%) |
| **Educational Level** |  |
| No Level | 708 (1.28%) |
| Primary | 10,334 (18.72%) |
| Secondary | 36,399 (65.95%) |
| Superior | 7,749 (14.04%) |
| **Civil status** |  |
| Single | 7,889 (27.92%) |
| With a partner | 20,361 (72.08%) |
| **Natural region** |  |
| Metropolitan Lima | 14,882 (26.96%) |
| Rest of coast | 15,651 (28.36%) |
| Mountain Range | 9,420 (17.07%) |
| Jungle | 15,237 (27.61%) |
| **Area of residence** |  |
| Urbana | 33,322 (73.84%) |
| Rural | 11,806 (26.16%) |
| **Mother's BMI** |  |
| Normal Weight | 20,991 (38.05%) |
| Overweight | 21,453 (38.88%) |
| Obesity | 12,729 (23.07%) |
| **Television Watching** |  |
| Never | 3,374 (7.35%) |
| Occasionally | 15,374 (33.48%) |
| Frequently | 27,174 (59.18%) |
| **Anemia in Mother** |  |
| No | 42,995 (78.52%) |
| Yes | 11,762 (21.48%) |
| **Mother's Smoking Activity** |  |
| Non-Smoker | 53,896 (97.65%) |
| Smoker | 1,294 (2.35%) |
| **Child's Age** |  |
| Less two years | 34,510 (75.14%) |
| 3 - 5 years | 11,420 (24.86%) |
| **Child's Gender** |  |
| Female | 27,094 (49.09%) |
| Male | 28,096 (50.91%) |
| **Child's Nutritional Status** |  |
| Normal Weight | 50,186 (90.93%) |
| Overweight | 3,908 (7.08%) |
| Obesity | 1,096 (1.99%) |

Figure 1 illustrates the relationships between various maternal factors and child BMI. In the upper left panel, a clear positive correlation is observed between the mother’s BMI and that of the child, visually supporting our finding that maternal obesity is significantly associated with an increased risk of childhood obesity. This trend is evidenced by the concentration of points ascending from left to right, indicating that child BMI also tends to increase as maternal BMI increases. On the other hand, the different panels of the graph show less pronounced relationships. The upper right panel presents the distribution of child BMI according to maternal television use levels, with box plots showing similar distributions for the three levels. Similarly, the lower left panel shows comparable distributions of child BMI between smoking and non-smoking mothers, supporting the absence of a significant association between maternal smoking and childhood obesity. Lastly, the lower right panel exhibits the relationship between maternal hemoglobin levels and child BMI, where no clear trend is observed.

**Figure 1.**
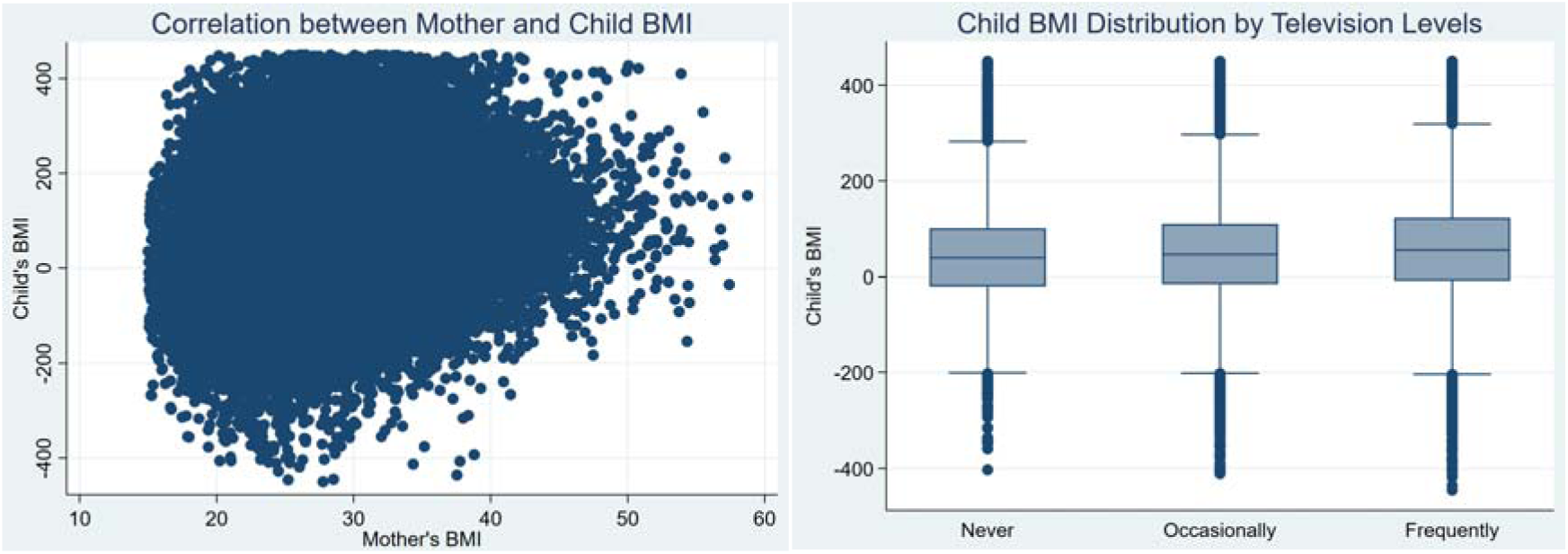
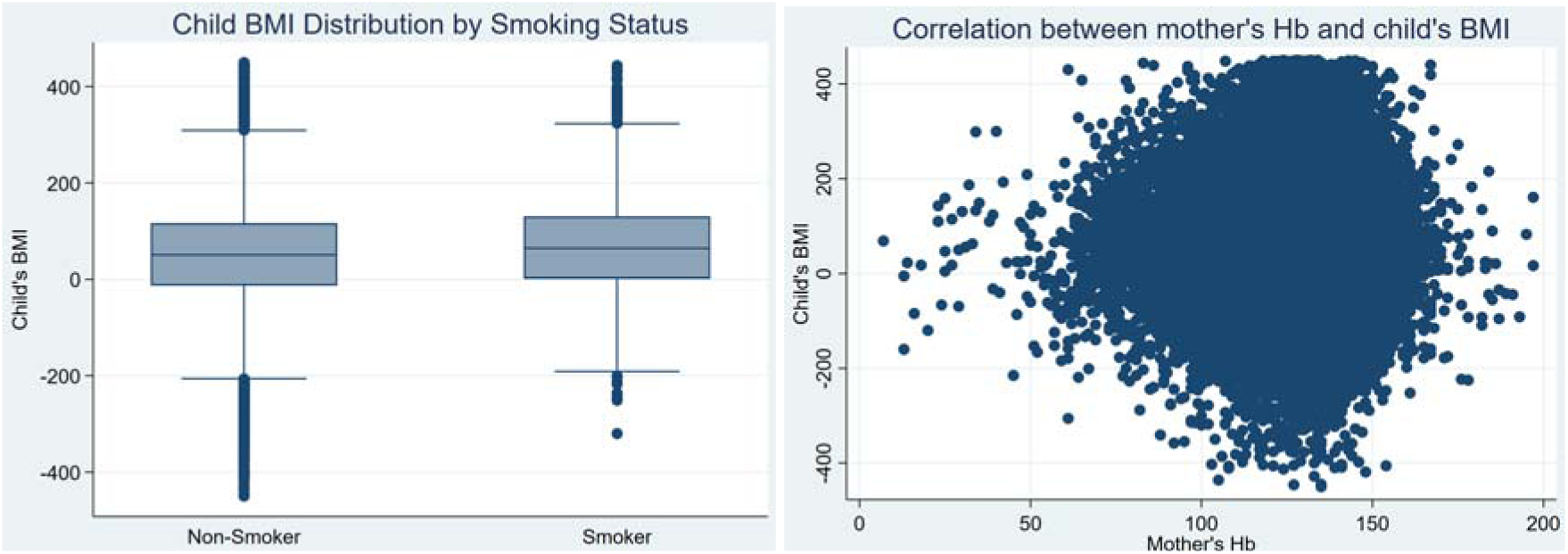
Associations between maternal BMI, television use, smoking, and maternal hemoglobin levels with child BMI.

Regarding the multivariate analysis, it can be observed that mothers with obesity are 1.52 times more likely to have children with obesity compared to mothers without this condition (aPR=1.52, 95% CI 1.40-1.65; p<0.001). However, no significant association was found between television use, the presence of maternal anemia, and maternal smoking status.

**Table 2.** Poisson regression analysis on the obesogenic environment and maternal obesity with childhood obesity.

| Characteristic | Univariable |  |  | Multivariable |  |  |
| --- | --- | --- | --- | --- | --- | --- |
|  | cPR | 95% CI | p-value | aPR* | 95% CI | p-value |
| **Obesity in Mother** |  |  |  |  |  |  |
| No Obesity | — | — |  | — | — |  |
| Obesity | 1.81 | 1.74, 1.89 | **<0.001** | 1.52 | 1.40, 1.65 | **<0.001** |
| **Television Watching** |  |  |  |  |  |  |
| Never | — | — |  | — | — |  |
| Occasionally | 1.32 | 1.20, 1.46 | **<0.001** | 1.06 | 0.86, 1.32 | 0.569 |
| Frequently | 1.74 | 1.58, 1.92 | **<0.001** | 1.17 | 0.96, 1.46 | 0.131 |
| **Anemia in Mother** |  |  |  |  |  |  |
| No | — | — |  | — | — |  |
| Yes | 0.93 | 0.89, 0.98 | **0.005** | 1.03 | 0.94, 1.13 | 0.534 |
| **Mother's Smoking Activity** |  |  |  |  |  |  |
| Non-Smoker | — | — |  | — | — |  |
| Smoker | 1.33 | 1.17, 1.51 | **<0.001** | 1.02 | 0.81, 1.26 | 0.889 |

\*Each factor has been independently adjusted for age, natural region, area of residence, educational level, wealth index, marital status, age, and sex of the child.
cRP: Crude prevalence ratio, aRP: Adjusted prevalence ratio.
95% CI: 95% confidence interval
Source: self-made

## Discussion

### Prevalence and Trends of Obesity in Mothers and Children

The findings of this study indicate that the prevalence of obesity in participating mothers and children under five years was 23.07% and 1.99%, respectively. These results are of public health importance as maternal obesity is associated with an increased risk of various health complications for both the mother and her offspring. Comparing these results with previous studies, a similar prevalence was found in other regions of Latin America. For example, the 2022 National Health and Nutrition Survey of Mexico (ENSANUT) reported an obesity prevalence of about 30% in women of childbearing age ^(13)^. These results coincide in low- and middle-income countries, and various factors contribute to these high prevalences, including changes in diet and physical activity patterns. However, it is also a significant problem in developed countries due to consuming processed foods and fats and increased urbanization involving lifestyle changes ^(14)^. Social and cultural factors could also affect women’s eating behaviors; being overweight may be considered a sign of health and prosperity, reducing women’s motivation to maintain a healthy weight ^(15)^.

On the other hand, regarding the prevalence of obesity in children under five years, the result of this study is consistent with that reported in a study conducted in Colombia, where they reported an obesity prevalence of 2.1% ^(16)^. While the prevalence of obesity in children is lower than in adults, childhood obesity is a critical indicator associated with long-term health risks, including the development of chronic diseases such as type 2 diabetes mellitus and cardiovascular diseases such as hypertension ^(17)^. One of the main reasons contributing to this prevalence is the access and availability of processed foods and fats, as they are more attractive to children due to their taste and mass marketing than healthy foods ^(18)^. Furthermore, urbanization and increased time spent on sedentary activities such as television use and video games have significantly reduced the time children invest in regular physical activity, in addition to the lack of safe spaces for recreational activities and food insecurity, which can increase the tendency towards the behaviors above ^(19)^.

Finally, it can be observed that in the trend analysis of maternal and child obesity prevalences in Peru, there was a significant increase in 2020; these results are mainly related to the impact of the COVID-19 pandemic. During this stage, families experienced drastic changes in everyday lifestyles, including less physical activity due to confinement restrictions and the closure of public spaces ^(20)^. Another factor to consider during the pandemic is the psychological stress generated by the fear of contracting the infection, which may have led to increased consumption of unhealthy foods to cope with this situation. The economic crisis resulting from the pandemic severely affected many families, reducing their ability to acquire nutritious foods ^(21)^.

### Maternal Obesogenic Environment

Regarding the obesogenic environment of participating mothers, the findings show that 59.18% use television frequently, 2.35% are smokers, and 21.48% have anemia. The high percentage of television use is consistent with other studies that have reported an association between excessive screen time and obesity. For example, one study found that women who spent more than three hours a day watching television had a higher body mass index and greater abdominal circumference than those who watched less television ^(22)^. It is known that television not only decreases the time available for physical activities but is also associated with the consumption of unhealthy foods due to exposure to advertising for food products of low nutritional quality ^(23)^.

Meanwhile, this study shows that only 2.35% of mothers are smokers, a relatively low result compared to other countries. In Spain, a study found that approximately 18% of women of reproductive age were smokers ^(24)^. However, smoking is associated with complications during pregnancy and can negatively affect fetal growth and proper child development.

Finally, the prevalence of anemia in our study is worrying and is consistent with results from other studies conducted in developing countries. One study reported an anemia prevalence of 53.1% in women of reproductive age, which is an indicator of poor nutrition and nutritional deficiencies, which can also contribute to the development of obesity ^(25)^. It should be taken into account that before these three dimensions, health policies and programs should be strengthened, considering that the obesogenic environment not only directly affects the mother but also affects the offspring immediately in health in an integral manner.

### Maternal Smoking and Childhood Obesity

In our study, the prevalence of smoking among mothers was relatively low, with only 2.35% of participants reporting being smokers. This figure is considerably lower than rates reported in other countries. For example, in Spain, a study found that approximately 18% of women of reproductive age were smokers ^(26)^. Despite this low prevalence, it is essential to consider the potential impact of maternal smoking on childhood obesity.

Although our multivariate analysis did not show a significant association between maternal smoking and childhood obesity after adjusting for multiple factors, existing literature suggests that this relationship deserves attention. A meta-analysis conducted by Oken et al. found that children whose mothers smoked during pregnancy were 50% more likely to be obese ^(27)^. More recent studies have supported this finding. For example, Møller et al. conducted a prospective cohort study that showed a significant increase in BMI and risk of overweight in children of mothers who smoked during pregnancy ^(28)^.

The proposed mechanisms to explain this association are diverse. It has been suggested that maternal smoking may alter fetal metabolism and cause epigenetic changes that predispose to obesity. Additionally, smoking during pregnancy has been associated with low birth weight, which paradoxically can increase the risk of later obesity due to accelerated compensatory growth ^(29)^.

It is important to note that our study’s lack of significant association could be due to several factors. The low smoking prevalence in our sample may have limited the statistical power to detect an association. Furthermore, our study was based on current smoking, while many studies focus on smoking during pregnancy, which could explain the differences in results.

Despite our findings, the accumulated evidence suggests that maternal smoking should continue to be considered as a potential risk factor for childhood obesity. Public health interventions aimed at reducing smoking in women of reproductive age could have additional benefits in preventing childhood obesity.

### Sedentary Behavior and Childhood Obesity

Our study evaluated maternal sedentary behavior through television use, finding that 59.18% of mothers reported watching television frequently. This high percentage of frequent television use is concerning, considering the growing evidence linking parental sedentary behavior with the risk of childhood obesity.

Although our multivariate analysis did not show a significant association between mothers’ frequent television use and childhood obesity after adjusting for multiple factors, existing literature suggests this relationship deserves careful consideration. Various studies have found significant associations between parents’ screen time and the risk of obesity in children.

For example, a longitudinal study conducted by Ishii et al. found that parents’ screen time was positively associated with children’s screen time and, consequently, a higher risk of childhood overweight and obesity ^(30)^. This finding underscores the importance of considering parents’ sedentary behaviors as contributing to the family’s obesogenic environment.

The mechanism by which frequent television use by mothers can influence childhood obesity is multifaceted. First, parents’ behavior is a model for children, so mothers who spend a lot of time watching television may inadvertently promote similar sedentary habits in their children. Additionally, watching TV reduces physical activity opportunities for mothers and children ^(31)^.

It is important to note that our study’s lack of significant association could be due to several factors. On one hand, our measure of television use might not fully capture sedentary behavior, as it does not include other forms of screen time, such as smartphone or computer use. Additionally, the impact of television use on childhood obesity may be mediated by other factors that we could not measure directly, such as family eating patterns or sleep quality ^(32)^.

Despite our findings, the accumulated evidence suggests that frequent television use and other sedentary behaviors of parents should continue to be considered as potential risk factors for childhood obesity. Future interventions to prevent childhood obesity should consider strategies to reduce screen time for the entire family, promoting alternative activities that foster a more active lifestyle.

### Maternal Anemia and Childhood Obesity

In our study, we found that 21.48% of mothers had anemia. This prevalence is concerning and consistent with other studies conducted in developing countries. For example, a global study estimated that the prevalence of anemia in women of reproductive age in low- and middle-income countries ranges between 20% and 40% ^(33)^.

Although our multivariate analysis did not show a significant association between maternal anemia and childhood obesity after adjusting for multiple factors, the literature suggests that this relationship is complex and deserves careful consideration. Maternal anemia and childhood obesity may be interconnected through several mechanisms, reflecting the complexity of the nutritional environment in which children develop.

A cohort study conducted by Brion et al. found that maternal anemia during pregnancy was associated with an increased risk of overweight in children at seven years of age ^(34)^. The authors suggested this could be due to altered fetal programming in response to maternal iron deficiency. On the other hand, maternal anemia can indicate a diet low in nutritional quality but high in empty calories, which could contribute to the family’s obesogenic environment. A study by Eckhardt et al. found a paradoxical coexistence of anemia and overweight in women from countries in nutritional transition, suggesting that both conditions may be the result of unbalanced diets ^(9)^.

Furthermore, maternal anemia can affect breastfeeding and infant feeding practices. A study by Zhao et al. found that mothers with anemia were more likely to introduce complementary foods early, which has been associated with an increased risk of childhood obesity ^(35)^.

It is important to note that our study’s lack of significant association could be due to several factors. On one hand, anemia may have different effects depending on when it occurs (for example, during pregnancy versus after childbirth). Additionally, the impact of maternal anemia on childhood obesity could be mediated by other factors that we could not measure directly, such as the quality of the family diet or infant feeding practices ^(36)^.

Despite our findings, the accumulated evidence suggests that maternal anemia should continue to be considered a relevant factor in the family obesogenic environment. Future interventions to prevent childhood obesity should consider comprehensive strategies that address maternal anemia and the nutritional quality of the family diet.

### Relationship Between Maternal Obesity and Childhood Obesity

This study revealed that mothers with obesity are 1.52 times more likely to have children with obesity compared to mothers who do not have this condition. These results are consistent with evidence highlighting a strong association between maternal obesity and childhood obesity. A study conducted in the United States found that children of mothers with obesity had a significantly higher risk of developing obesity, relating these results to genetic factors and shared behaviors or attitudes ^(37)^. Maternal obesity can influence fetal programming through different epigenetic mechanisms and can also shape eating and physical activity habits in children, which favors the perpetuation of the intergenerational obesity cycle ^(38)^.

A combination of genetic, biological, and environmental factors could explain the relationship between maternal obesity and childhood obesity. Genetically, children of mothers with obesity can inherit a predisposition to weight gain and abdominal fat accumulation, which can influence the child’s metabolism and appetite regulation and increase the risk of developing obesity (39). On the other hand, maternal obesity can affect the intrauterine environment, leading to epigenetic changes that can predispose the child to obesity.

These changes may include alterations in the expression of genes related to glucose metabolism and fat accumulation, affecting birth weight and subsequent growth development of the child ^(40)^. Furthermore, mothers with obesity may transmit unhealthy eating habits and lifestyles to their children ^(41)^. Eating patterns and physical activity observed at home are models that children tend to imitate. Additionally, maternal stress and depression, which are more prevalent in women with obesity, can affect parenting practices and the mother’s ability to foster healthy lifestyles in her offspring. These factors create an obesogenic environment that increases the likelihood of children developing obesity during their growth ^(42)^.

Finally, an important influencing factor is also the socioeconomic environment. This variable has been related to maternal obesity; consequently, mothers with obesity may have less access to resources to maintain a healthy diet and active lifestyle due to economic limitations ^(43)^. Food insecurity is more common in families with lower incomes, which can lead to the purchase of cheap and calorically dense foods that are less nutritious, favoring the development of obesity in their children ^(44)^.

### Strengths and Limitations of the Study

This study presents several significant strengths. Firstly, it is based on data from DHS, a nationally representative survey, which provides a large and diverse sample that allows for generalizing results at the national level. Additionally, the study incorporates a wide range of variables from the maternal obesogenic environment, including not only maternal obesity but also factors such as television use, smoking, and anemia, providing a more comprehensive view of the family environment. The multivariate analysis, adjusted for numerous potential confounding factors, strengthens the validity of the findings and allows for a more precise interpretation of the observed associations.

However, this study also presents some limitations that should be considered. Firstly, being a cross-sectional study, it is impossible to establish direct causal relationships between factors of the maternal obesogenic environment and childhood obesity, so the approach is still exploratory. The self-reported nature of some variables, such as television use and smoking, may introduce information biases. Furthermore, although several aspects of the obesogenic environment were included, it was impossible to directly measure essential factors such as detailed family diet or physical activity levels, which could have provided a more complete picture. Finally, although adjustments were made for multiple confounding factors, residual confounding is always possible by unmeasured or unincluded variables in the analysis.

## Conclusions

A significant association was found between maternal obesity and the presence of obesity in children under five years of age. Furthermore, although no significant associations were found between frequent television use, smoking, and maternal anemia with childhood obesity, these elements that are part of the obesogenic environment remain relevant in the broader context of family health. On the other hand, the prevalence of maternal obesity (23.07%) and childhood obesity (1.99%) underscores the importance of addressing this public health problem. These findings highlight the complexity of the obesogenic environment and the need to consider multiple factors in preventing childhood obesity in Peru.

Based on the results of this study, the development and implementation of comprehensive interventions that address multiple aspects of the family obesogenic environment are recommended. These interventions should promote healthy lifestyles for the entire family, including improving eating habits, increasing physical activity, and reducing sedentary time. It is suggested that maternal and child health programs be strengthened, strategies to prevent and manage maternal obesity should be incorporated, maternal nutrition should be improved in general, and problems such as anemia should be addressed. In addition, it is recommended that public policies be implemented that promote healthy environments, such as regulating the advertising of unhealthy foods aimed at children and creating safe spaces for physical activity. Future research should further explore the mechanisms by which the maternal obesogenic environment influences childhood obesity, including longitudinal studies that allow for establishing causal relationships and evaluating the effectiveness of the proposed interventions.

## Authors’ contribution

Joan A. Loayza-Castro: Conceptualization, Investigation, Methodology, Resources, Writing - Original Draft, Writing - Review & Editing

Luisa Erika Milagros Vásquez-Romero: Investigation, Project administration, Writing - Original Draft, Writing - Review & Editing

Fiorella E. Zuzunaga-Montoya: Investigation, Resources, Writing - Original Draft, Writing - Review & Editing

Jhonatan Roberto Astucuri Hidalgo: Software, Data Curation, Formal analysis, Writing - Review & Editing

Nataly Mayely Sanchez-Tamay: Validation, Visualization, Writing - Original Draft, Writing - Review & Editing

Víctor Juan Vera-Ponce: Methodology, Supervision, Funding acquisition, Writing - Review & Editing

## Acknowledgments

A special thanks to the members of Universidad Nacional Toribio Rodríguez de Mendoza de Amazonas (UNTRM), Amazonas, Peru for their support and contributions throughout the completion of this research.

## Informed consent

The primary study from which the database was obtained provided the required informed consent, however, for the present study it was not required.

## Data availability

The data supporting the findings of this study can be accessed by the original research paper
at the follow link: https://proyectos.inei.gob.pe/microdatos/

